# Utilisation of laboratory test results for patient management by clinicians at two large referral hospitals in Zambia

**DOI:** 10.1101/2020.09.23.20200071

**Authors:** Sabe Mwape, Victor Daka, Scott K. Matafwali, Kapambwe Mwape, Jay Sikalima, Pipina A. Vlahakis, Memory C. Kalolekesha, Namaunga K. Chisompola, Victor Chalwe

**Affiliations:** University of Lusaka, P.O Box 37413, Lusaka, Zambia; Catholic Relief Services, P.O Box 38086, Lusaka, Zambia; Copperbelt University, School of Medicine, Basic Science Department, P.O Box 71191 Ndola, Zambia; Copperbelt University, School of Medicine, Department of Clinical Sciences, Public Health Unit, P.O Box 71191 Ndola, Zambia; National Health Research Authority, P.O Box 30075, Lusaka, Zambia; Tropical Diseases Research Centre, P.O Box 71769, Ndola, Zambia

**Keywords:** utilisation, clinicians, laboratory tests, Ndola Teaching Hospital, Arthur Davison Childrens Hospital

## Abstract

**Background:** Medical laboratory diagnosis is a critical component of patient management in the healthcare setup. Despite the availability of laboratory tests, clinicians may not utilise them to make clinical decisions. We investigated utilsation of laboratory tests for patient management among clinicians at Ndola Teaching Hospital (NTH) and Arthur Davison Childrens Hospital (ADCH), two large referral hospitals in the Copperbelt Province, Ndola, Zambia.

**Method:** We conducted a descriptive cross-sectional study among clinicians. The study deployed self-administered questionnaires to evaluate clinician utilisation, querying and confidence in laboratory results. Additional data on demographics and possible laboratory improvements were also obtained. Data were entered in Microsoft excel and exported to SPSS version 16 for statistical analysis.

**Results:** Of the 80 clinicians interviewed, 96.2% (77) reported using laboratory tests and their results in patient management. 77.5% (62) of the clinicians indicated they always used laboratory results to influence their patient management decisions. Of the selected laboratory tests, clinicians were more confident in using haemoglobin test results (91.2%). There was no statistically significant association between the clinicians gender or qualification and use of test results in patient management.

**Conclusion:** Our findings show that despite the majority querying laboratory results, most of the clinicians use laboratory results for patient management. There is need for interactions between the laboratory and clinical area to assure clinician confidence in laboratory results.

## 1.0 Introduction

Clinicians are tasked with making informed decisions regarding patient care based on available clinical information. This can be derived from patient history and physical examination [1]. Although data elicited from the history and physical examination may be sufficient for making a diagnosis or for guiding therapy, more information is often required. This information can be obtained from diagnostic tests such as medical laboratory tests (MLTs) [2,3].

Medical laboratory tests greatly influence health care decisions and are among the least expensive components of the health care pathway [4]. These tests provide objective information about a person’s health which can be used for many purposes [4,5]. One such purpose is to decrease diagnostic uncertainty in a patient presenting with non-specific signs and symptoms as well as to monitor treatment response in a patient with a prior diagnosis after commencing treatment [5]. MLTs may also be requested for screening or case-finding, and in some cases, at a patient’s request [5].

Accessibility to MLTs varies with some tests only available as point-of-care while others may only be available at a reference laboratory [6,7]. This greatly impacts patient management. Clinical studies have demonstrated instances in which diagnostic tests were used for patient care and how these tests influenced patient care [8,9]. The studies further stressed the benefits of using medical laboratory tests in health services and emphasized the critical role the laboratory results play in effective patient management [10]. Additionally, MLTs have demonstrated to reduce treatment costs due to empiric therapy as well as prolonged hospital stays [8].

Despite the highlighted benefits, clinicians often do not use laboratory tests in patient management [11]. A trend of low utilization of laboratory results in patient management has been observed as one of the major factors negatively impacting outcomes for most patients accessing health care services in resource-constrained countries of Sub-Saharan Africa [11,12]. Coupled with the challenges of high disease burden, scarce funding, inadequate health personnel, poor health linkages, and inadequate infrastructure, the practice has the potential to negatively affect the provision of quality health services [13]. We, therefore, undertook a cross-sectional descriptive study of the utilisation of laboratory services by clinicians at NTH and ADCH, two large tertiary referral hospitals in Ndola, Zambia.

## 2.0 Methods

### 2.1 Study Site

The study was conducted at NTH and ADCH in Ndola, Zambia. NTH is a proveincial referral hospital for the northern region of Zambia which includes Copperbelt, Luapula and North-Western Provinces of Zambia [14]. ADCH, the only paedaetric hospital in Zambia, is located approximately 2km from the city center of Ndola with a bed capacity of 250 [15]. The two hospitals are approximately 2km apart and with inherent similarities in staffing as clinicians may rotate between the two hospitals.

### 2.2 Target population

The target population was clinicians working at NTH and ADCH. Primary data were collected from 80 practicing clinicians attending to patients at the two hospitals.

### 2.3 Sampling

All available clinicians working at the two hospitals during the study period and willing to participate in the study were enrolled.

### 2.4 Data collection

Data were collected using a self-administered questionnaire. Trained research assistants were available for the clinician to consult where they needed clarifications. The questionnaire was divided into questions on demographics, utilisation of laboratory results, querying of results, confidence in laboratory tests and clinician’s perception on laboratory needs and improvements. Utilisation and querying of results were assessed through a three point scale–yes, sometimes and no.

### 2.5 Data analysis

Data were entered in excel and cleaned for consistency and range checks and then exported to SPSS version 16 for statistical analysis. We conducted an internal reliability test for our questionnaire using the Cronbach’s alpha according to methods described by Taber [16] and obtained an alpha value of 0.631 indicating acceptable reliability. The chi-square test was used to determine factors associated with the use of laboratory tests and a p-value less than 0.05 (p < 0.5) was considered significant.

## 3.0 Ethical considerations

Ethical clearance was granted by ERES Converge Institutional Review Board (IRB) and authority to conduct the study was obtained from the National Health Research Authority (NHRA). Written permission to carry out the study was sought from the Provincial Medical Office for Copperbelt Province. All respondents that accepted to be part of the study were provided with informed consent and were told the study objectives, procedure, risks, and benefits, and participant’s rights. No identifiers were used to ensure confidentiality and anonymity of respondents. Access to all study materials including questionnaires was restricted to the investigator and assistants.

## 4.0 Results

There were more male respondents (n=51, 64%) than females. Two-thirds of the respondents were aged between 20 and 39 years of age had been one to three years in service. Majority of the respondents were medical doctors (86.3%) and were trained in Zambia (Table 1).

**Table 1.**
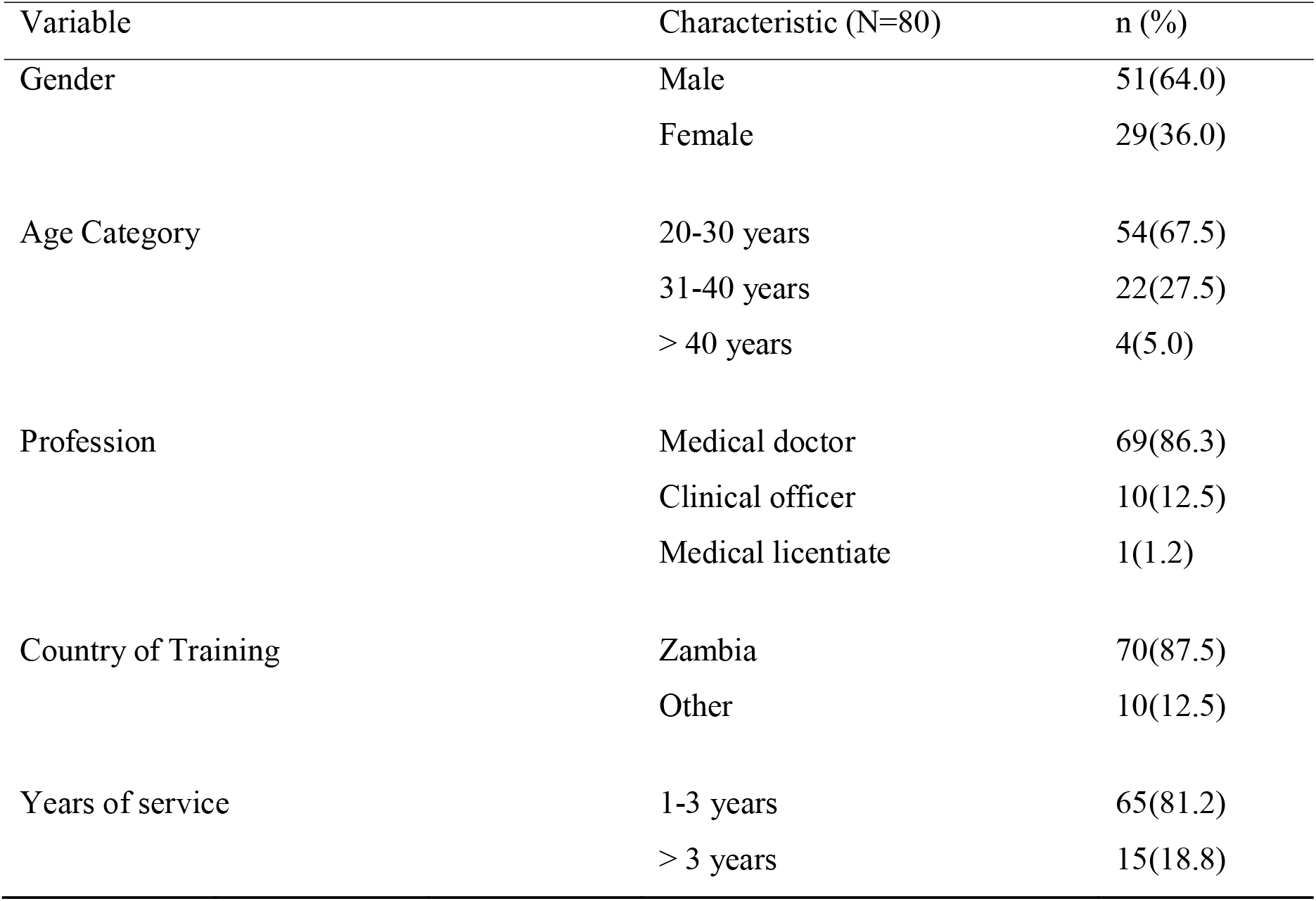
Demographic characteristics of respondents

| Variable | Characteristic (N=80) | n (%) |
| --- | --- | --- |
| Gender | Male | 51(64.0) |
|  | Female | 29(36.0) |
| Age Category | 20-30 years | 54(67.5) |
|  | 31-40 years | 22(27.5) |
|  | > 40 years | 4(5.0) |
| Profession | Medical doctor | 69(86.3) |
|  | Clinical officer | 10(12.5) |
|  | Medical licentiate | 1(1.2) |
| Country of Training | Zambia | 70(87.5) |
|  | Other | 10(12.5) |
| Years of service | 1-3 years | 65(81.2) |
|  | > 3 years | 15(18.8) |

Table 2 summarises the use, querying and clinician request for repeat results. The majority of the respondents reported using laboratory results all the time (62, 77.5%). Almost half of the respondents reported querying laboratory results all the time (39, 48.8%). There were more clinicians who reported never requesting to repeat a tuberculosis (TB) test (50, 62.5%) and microscopy culture and sensitivity result (47, 58.8%). There were more requests for a repeat malaria result (50, 62.5%)

**Table 2.** Use, querying of results and frequency of repeating tests

| Variable | All the time n(%) | Some of the time n(%) | Never n(%) |
| --- | --- | --- | --- |
| Use of requested laboratory results | 62 (77.5) | 15(18.7) | 3(3.8) |
| Querying of laboratory test results | 39(48.8) | 34(42.5) | 7(8.8) |
| Requested repeat of test/ Tests requested for repeat |  |  |  |
| AST/ALT | 11(13.8) | 33(41.2) | 36(45.0) |
| CD4 | 16(20) | 46(57.5) | 18(22.5) |
| Hb | 35(43.8) | 38(47.5) | 7(8.8) |
| Malaria | 7(8.8) | 50(62.5) | 23(28.8) |
| TB | 1(1.2) | 29(36.2) | 50(62.5) |
| Microscopy, Culture and Sensitivity | 3(3.8) | 30(37.5) | 47(58.8) |

Table 3 shows the confidence of clinicians in laboratory results. Respondents reported the highest confidence in haemoglobin (Hb) results (73, 91.2%) and CD4 results (72, 90%) while slightly over one-third of the respondents reported low confidence in malaria results (31, 38.8%).

**Table 3.** Confidence in laboratory results by test performed

| Variables | Yes |  | No |  |
| --- | --- | --- | --- | --- |
|  | n | (%) | n | (%) |
| AST/ALT | 69 | 86.2 | 11 | 13.8 |
| CD4 | 72 | 90 | 8 | 10 |
| Hb | 73 | 91.2 | 7 | 8.8 |
| Malaria | 31 | 38.8 | 49 | 60 |
| TB | 51 | 63.8 | 29 | 36.2 |
| MCS | 60 | 75 | 18 | 22.5 |

Table 4 shows the most compelling reasons given for repeating a test before confirming a diagnosis, monitoring and follow up. Notably, heamoglobin tests were also repeated for purposes of dissatisfaction (23, 28.8%) and querying (50, 62.5%).

**Table 4.** Reasons for repeating test

| Variables | AST/ALT |  | CD<sub>4</sub> |  | HB |  | Malaria |  | TB |  | MCS |  |
| --- | --- | --- | --- | --- | --- | --- | --- | --- | --- | --- | --- | --- |
|  | Freq | (%) | Freq | (%) | Freq | (%) | Freq | (%) | Freq | (%) | Freq | (%) |
| Diagnosis | 45 | 56.2 | 4 | 5 | 5 | 6.2 | 43 | 53.8 | 43 | 53.8 | 51 | 63.8 |
| Monitoring | 20 | 25 | 67 | 83.8 | 42 | 52.5 | 9 | 11.2 | 21 | 26.2 | 23 | 28.7 |
| Follow up | 15 | 18.8 | 9 | 11.2 | 30 | 37.5 | 26 | 32.5 | 16 | 20 | 6 | 7.5 |
| Dissatisfied | - | - |  |  | 23 | 28.8 |  |  |  |  |  |  |
| Querying | - | - |  |  | 50 | 62.5 | 2 | 2.5 |  |  |  |  |
| Supportive | - | - |  |  | 2 | 2.5 |  |  |  |  |  |  |
| Treatment | - | - |  |  | 1 | 1.2 |  |  |  |  |  |  |
| Total | 80 | 100 | 80 | 100 | 80 | 100 | 80 | 100 | 80 | 100 | 80 | 100 |

Table 5 shows that most of the clinicians reported some interaction with laboratory staff either daily (34, 42.5%) or some of the times (38, 47.5%) with half of the respondents reporting having interacted with laboratory personnel as working colleagues (41, 51.2%).

**Table 5.**
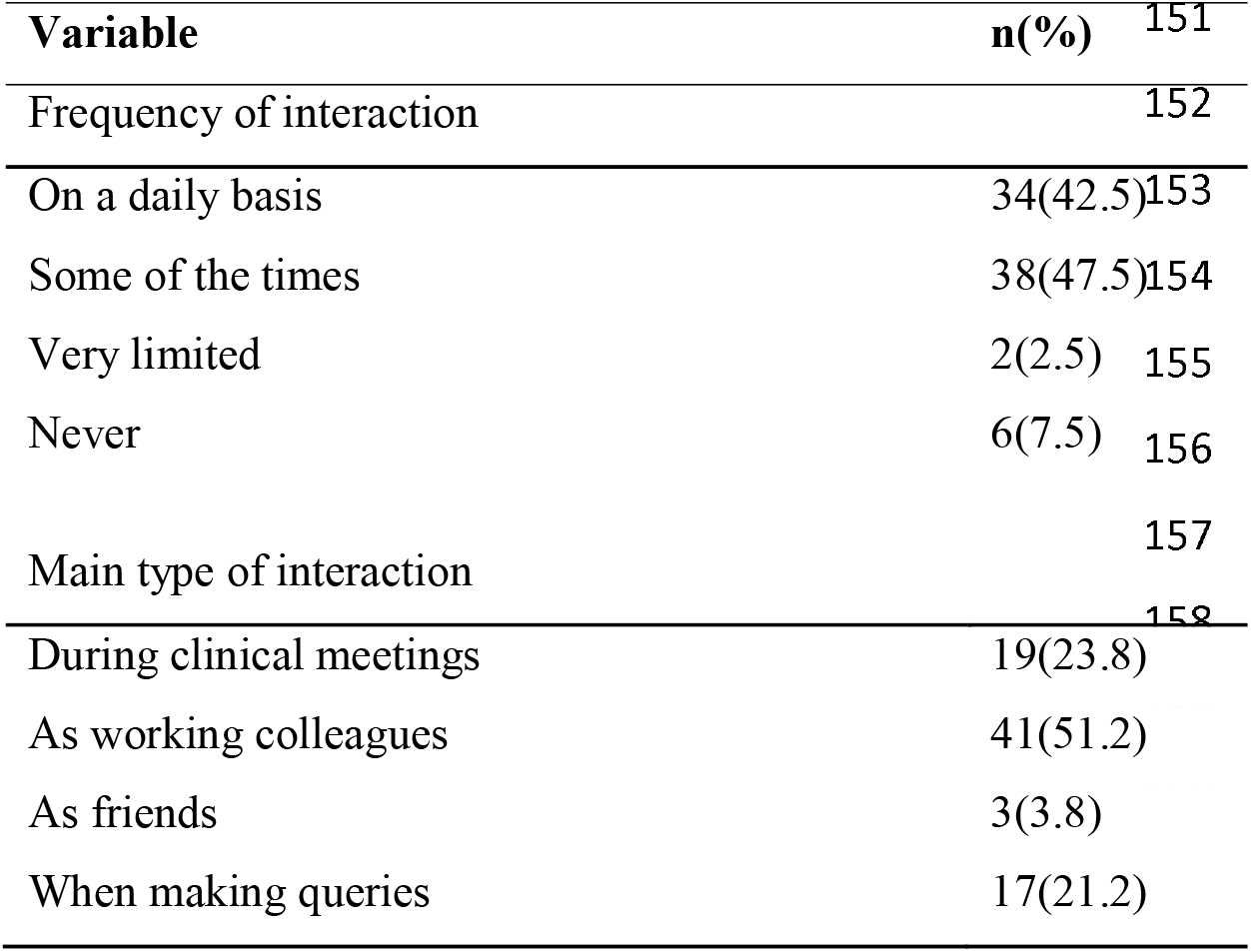
Frequency and type of interaction

**Table 6.**
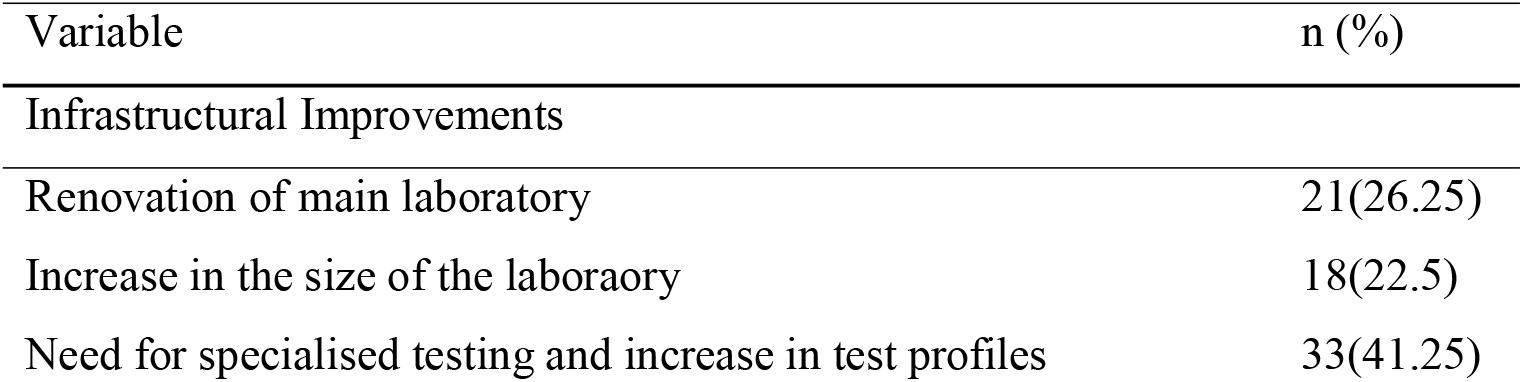
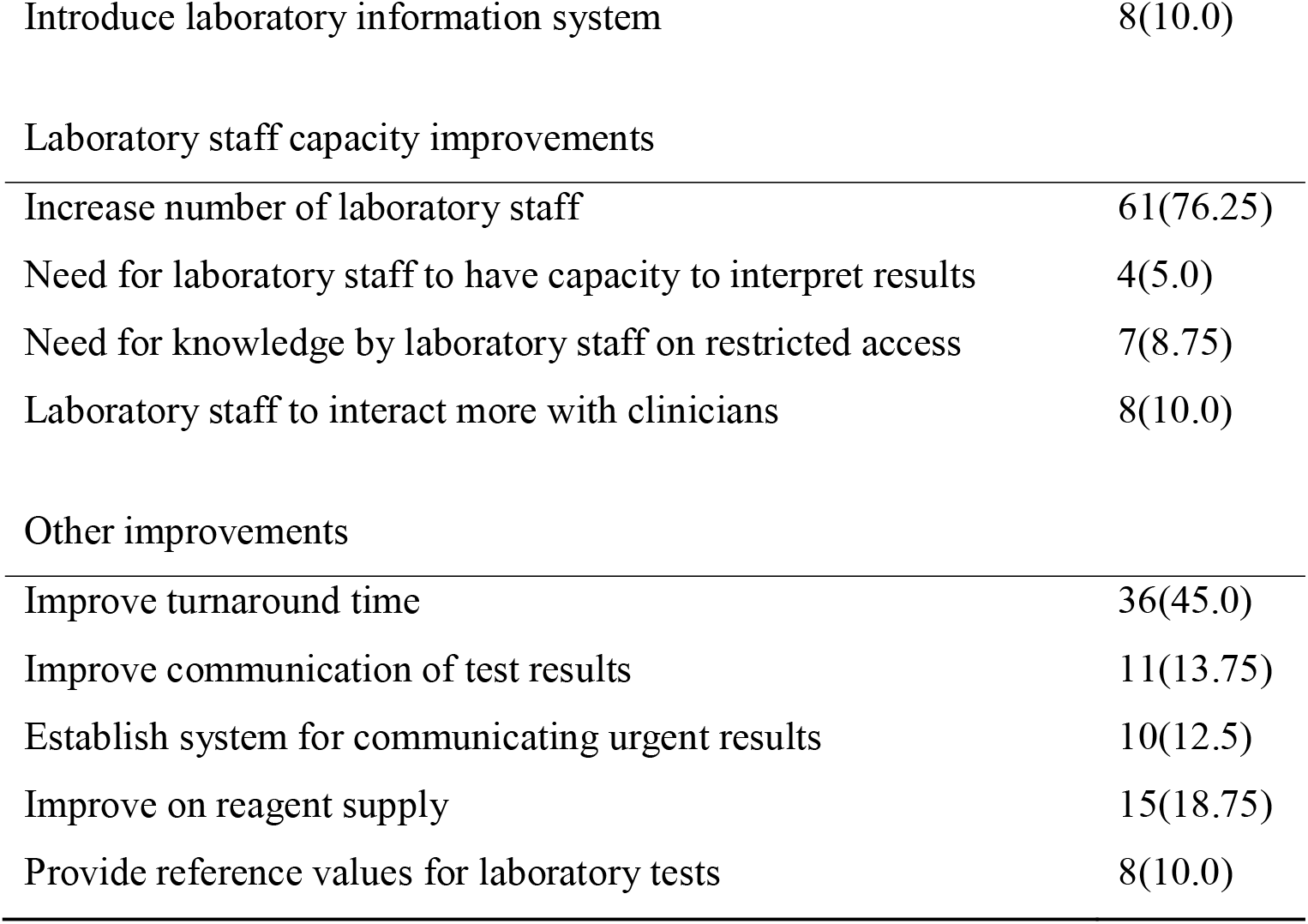
Clinicians’ perceptions on the required laboratory improvements

More of the clinicians felt there was need to improve test profiles including specialized testing (33, 41.25%). The majority of the respondents (61, 76.25%) felt the laboratory was understaffed and needed to recruit more staff to meet the testing demands. Close to half (36, 45,0%) of the clinicians reported that test results took long and there was need to reduce the turnaround time (TAT).

### Association of demographic variables and utilization of laboratory tests

There was no association between utilization of laboratory tests and gender (p=0.791) and years of practice (p=0.308).

## 5.0 Discussion

Our findings demonstrated that all of the clinicians ordered a laboratory test at one time during their interaction with patients, although not all the ordered tests were used for decision making in patient management. Our findings also show that female clinicians used laboratory test results more often than male clinicians in patient management. The reasons for this occurrence remain speculative given that the analysis for a likely association between clinicians’ gender and use of lab results in patient management showed that the two were not significantly related. However, it can be argued that like most studies with smaller sample sizes, increasing the sample size and frame would yield slightly different results; in this case a significant association between the two variables [17].

The study findings also showed that three quarters of the clinicians always used laboratory results of requested tests to influence their patient management decisions. These findings are higher than the those reported from Malawi in 2009 where 64% of clinicians were using laboratory results in patient management decisions [10]. However similar findings were reported in 2015 where 70% of clinicians in Malawi always used test results in patient management [11]. These differences could be due to differences in study populations and power in the two studies.

In our study we demonstrated that about half of clinicians often questioned laboratory results, this is much lower than the reported in a study by Moyo and others in Malawi who reported 70% of the clinicians questioned results [11]. The querying of results by clinicians often leads to requests for repeat of tests. Clinicians often argue that the clinical presentation of patients may not be consistent with the laboratory results [18]. A study in Ghana reported a high reliance on clinical symptoms in diagnosing malaria [19]. In this study, results for haemoglobin were the most repeated followed by CD4 count and malaria tests. Analogous results were found in a study by Bates and Adu [20], in which following a survey of 205 laboratories in Ghana located in regional and district government hospitals, Hb and malaria had the least accurate results prompting clinicians to repeat the tests. The main reason for requesting repeat tests in our study was to confirm a diagnosis, unlike a study in Ghana where the doubts regarding the accuracy of results was the main reason for repeating the test [8]. The findings in this study could be attributed to delays in receiving results (long TAT) as the most likely reason as almost half requested an improved TAT. Additionally, clinicians may also be more likely to believe automated methods rather than manual methods [21]. Therefore, there is a need to adopt strategies that reduce TATs to address the trend.

The findings showed that those who qualify with diplomas and are clinical officers and medical licentiates tend to use lab results more than the degree holders who are medical doctors in patient management. This could be due to the medical doctors having more confidence and relying on clinical experience than the others [22]. The study also found that clinicians who interact with laboratory staff tend to use laboratory results more than those who never. Clinicians’ interaction with laboratory staff was significantly associated with the use of laboratory results. This can be due to constant interaction between clinicians and laboratory staff creating an opportunity for improvements in service delivery [11].

In this study, clinicians suggested increasing the staffing in the hospital laboratory, an increase in test profile and platforms, the establishment of a system to communicate urgent results and ensuring an adequate supply of laboratory reagents and consumables as areas needing support for the improvement of laboratory services. Other areas included renovating the laboratories, increasing the size of the laboratory, restricting access to the laboratory and the need for more interaction between laboratory staff and clinicians. These suggestions are similar to those reported by clinicians in a study done by Kundai and others and Petti and others [11,23]. The shortage of both personnel and supplies in laboratories contributes to a lengthy turnaround time as reported in a study by Lia and others [24]. Thus, the decision to order laboratory tests is often a choice between accuracy and expediency. This can lead to diagnoses often being made using less reliable and less valid signs and symptoms.

## 6.0 Conclusion

Our study demonstrated that laboratory test results are used by clinicians in patient management at NTH and ADCH. We also found that a high number of clinicians query results and have little confidence in results for some tests such as malaria. Our findings present results from regional tertiary hospitals. We recommend further studies to understand the impact of underutilizing laboratory results in a clinical setting in other health facilities in Zambia.

## 7.0 Limitations

Due to the self-admisnisterd nature of our study, we cannot rule out possible responder bias. Utilisation of laboratory results by individual clinicians could also not be verified as investigation of clinical records were not in the scope of this study. Our study found no association between some selected demographic factors and utilisation of laboratory results. This could have been affected by the limited sample size. Further, our study was limited by the short sampling period that it was thereby not including clinicians or key informants who were not available during the period of study. This could have missed out individuals that could have provided valuable information possibly omitted by the sampled participants.

## Data Availability

The data used in this study is available from the corresponding author upon request

## 8.0 Acknowledgments

We acknowledge the staff and management at NTH and ADCH for facilitating this study.

## 9.0 Competing interests

The authors declare that they have no financial or personal relationship(s) that may have inappropriately influenced them in writing this article.

## 10.0 Author contributions

Conception, design and data collection was done by SM, JS, VC and VD. Data analysis, interpretation and manuscript writing was conducted by SM, SKM, MK, PAV, NKC, VD and KM. Supervision of the study was done by VC. All authors reviewed and approved the final version of the manuscript.

## Notes

### Competing Interest Statement

The authors have declared no competing interest.

### Funding Statement

No external funding was obtained for this study

### Author Declarations

ERES Converge ethics committee

## References

1. Papadakis M, McPhee S, Rabow M. Current Medical Diagnosis and Treatment 2020 ü Access Medicine ü McGraw-Hill Medical [Internet]. 2020 [cited 2020 Jun 11]. Available from: https://accessmedicine.mhmedical.com/book.aspx?bookID=2683

2. Hajjaj FM, Salek MS, Basra MKA, Finlay AY. Non-clinical influences on clinical decision-making: A major challenge to evidence-based practice. Vol. 103, Journal of the Royal Society of Medicine. Royal Society of Medicine Press; 2010. p. 178–87.

3. Wians FH. Clinical Laboratory Tests: Which, Why, and What Do The Results Mean? Lab Med [Internet]. 2009 Feb 1 [cited 2020 Aug 22];40(2):105–13. Available from: https://academic.oup.com/labmed/article-lookup/doi/10.1309/LM4O4L0HHUTWWUDD

4. Ngo A, Gandhi P, Miller WG. Frequency that Laboratory Tests Influence Medical Decisions. J Appl Lab Med An AACC Publ [Internet]. 2017 Jan 1 [cited 2020 Jun 11];1(4):410–4. Available from: https://academic.oup.com/jalm/article/1/4/410-414/5587412

5. Rohr UP, Binder C, Dieterle T, Giusti F, Messina CGM, Toerien E, Moch H, Hendrikschäfer H. The value of in vitro diagnostic testing in medical practice: A status report. PLoS One. 2016 Mar 1;11(3).

6. Nkengasong JN, Mesele T, Orloff S, Kebede Y, Fonjungo PN, Timperi R, Birx D. Critical role of developing national strategic plans as a guide to strengthen laboratory health systems in resource-poor settings. In: American Journal of Clinical Pathology. Am J Clin Pathol; 2009. p. 852–7.

7. Mazaba M, Mwaba P, Droti B, Kagulura S, Makasa C, Masaninga F, Kachimba J, Vwalika B, Mufunda J. Leveraging Existing Laboratory Capacity towards Universal Health Coverage: A Case of Zambian Laboratory Services. Med J Zambia. 2016;43(2):88–93.

8. Bates I, Maitland K. Are laboratory services coming of age in sub-Saharan Africa? Clin Infect Dis [Internet]. 2006 Feb 1 [cited 2020 Jun 11];42(3):383–4. Available from: http://www.ncbi.nlm.nih.gov/pubmed/16392085

9. Anonychuk A, Beastall G, Shorter S, Kloss-Wolf R, Neumann P. A Framework for Assessing the Value of Laboratory Diagnostics. Healthc Manag Forum [Internet]. 2012 Oct 1 [cited 2020 Jun 11];25(3_suppl):S4–11. Available from: http://journals.sagepub.com/doi/10.1016/j.hcmf.2012.07.015

10. Mepham SO, Squire SB, Chisuwo L, Kandulu J, Bates I. Utilisation of laboratory services by health workers in a district hospital in Malawi. J Clin Pathol. 2009 Oct;62(10):935–8.

11. Moyo K, Porter C, Chilima B, Mwenda R, Kabue M, Zungu L, Sarr A. Use of laboratory test results in patient management by clinicians in Malawi. Afr J Lab Med. 2015 May 13;4(1).

12. Okeke IN. Diagnostic insufficiency in Africa. Clin Infect Dis [Internet]. 2006 May 15 [cited 2020 Jun 11];42(10):1501–3. Available from: http://www.ncbi.nlm.nih.gov/pubmed/16619170

13. Hailu HA, Yalew A, Desale A, Asrat H, Kebede S, Dejene D, Abebe H, Gashu A, Moges B, Yemanebrhane N, Melese D, Ayele BT, Kebede A, Abate E. Physicians’ satisfaction with clinical laboratory services at public hospitals in Ethiopia: A national survey. PLoS One. 2020 Apr 1;15(4):e0232178.

14. Chanda W, Manyepa M, Chikwanda E, Daka V, Chileshe J, Tembo M, Kasongo J, Chipipa A, Handema R, Mulemena JA. Evaluation of antibiotic susceptibility patterns of pathogens isolated from routine laboratory specimens at Ndola Teaching Hospital: A retrospective study. Abuelo A, editor. PLoS One [Internet]. 2019 Dec 23 [cited 2020 Aug 28];14(12):e0226676. Available from: https://dx.plos.org/10.1371/journal.pone.0226676

15. Mwale K. Intestinal Infestations in Under-Five Children in Zambia. Int J MCH AIDS [Internet]. 2016 Jan 1 [cited 2020 Aug 28];4(2):40–6. Available from: https://europepmc.org/articles/PMC4948131

16. Taber KS. The Use of Cronbach’s Alpha When Developing and Reporting Research Instruments in Science Education. Res Sci Educ [Internet]. 2018 Dec 1 [cited 2020 Aug 22];48(6):1273–96. Available from: https://link.springer.com/article/10.1007/s11165-016-9602-2

17. Faber J, Fonseca LM. How sample size influences research outcomes. Dental Press J Orthod [Internet]. 2014 Jul 1 [cited 2020 Aug 28];19(4):27–9. Available from: /pmc/articles/PMC4296634/?report=abstract

18. Carter J, Östensen H, Heuck CC. Good clinical diagnostic practice A guide for clinicians in developing countries to the clinical diagnosis of disease and to making proper use of clinical diagnostic services. 2005.

19. Polage CR, Bedu-Addo G, Owusu-Ofori A, Frimpong E, Lloyd W, Zurcher E, Hale D Von, Petti CA. Laboratory use in Ghana: Physician perception and practice. Am J Trop Med Hyg. 2006;75(3):526–31.

20. Bates I, Bekoe V, Asamoa-Adu A. Improving the accuracy of malaria-related laboratory tests in Ghana [Internet]. Vol. 3, Malaria Journal. Malar J; 2004 [cited 2020 Jul 13]. Available from: https://pubmed.ncbi.nlm.nih.gov/15516269/

21. Lippi G, Da Rin G. Advantages and limitations of total laboratory automation: A personal overview [Internet]. Vol. 57, Clinical Chemistry and Laboratory Medicine. De Gruyter; 2019 [cited 2020 Sep 19]. p. 802–11. Available from: https://pubmed.ncbi.nlm.nih.gov/30710480/

22. Hecimovich MD, Volet SE. Importance of Building Confidence in Patient Communication and Clinical Skills Among Chiropractic Students. J Chiropr Educ [Internet]. 2009 Oct 1 [cited 2020 Aug 26];23(2):151–64. Available from:/pmc/articles/PMC2759993/?report=abstract

23. Petti CA, Polage CR, Quinn TC, Ronald AR, Sande MA. Laboratory Medicine in Africa: A Barrier to Effective Health Care. Clin Infect Dis. 2006 Feb 1;42(3):377–82.

24. Petrose LG, Fisher AM, Douglas GP, Terry MA, Muula A, Chawani MS, Limula H, Driessen J. Assessing perceived challenges to laboratory testing at a malawian referral hospital. Am J Trop Med Hyg. 2016 Jun 1;94(6):1426–32.

